# An expanded population of CD8^dim^ T cells with features of mitochondrial dysfunction and senescence is associated with persistent HIV-associated Kaposi’s sarcoma under ART

**DOI:** 10.1101/2022.02.23.22271244

**Authors:** Genevieve T Clutton, Ann Marie K Weideman, Nilu P Goonetilleke, Toby Maurer

**Affiliations:** Department of Microbiology & Immunology, UNC Chapel Hill School of Medicine, Chapel Hill, North Carolina 27599, USA; Department of Biostatistics and Center for AIDS Research, UNC Chapel Hill, Chapel Hill, North Carolina 27599, USA; UNC HIV Cure Center, UNC Institute of Global Health and Infectious Diseases, Chapel Hill, North Carolina 27599, USA; Department of Dermatology, Indiana University, Indiana, USA

**Keywords:** T cells, CD8 co-receptor, KSHV, Kaposi’s sarcoma, Tumor immunity, HIV, Metabolism, Senescence, Mitochondria, MitoTracker, PGC-1α, Proliferation

## Abstract

HIV-associated Kaposi’s sarcoma (KS), which is caused by Kaposi’s sarcoma-associated herpesvirus, usually arises in the context of uncontrolled HIV replication and immunosuppression. However, disease occasionally persists in individuals with durable HIV viral suppression and CD4 T cell recovery under anti-retroviral therapy (ART). The underlying mechanisms associated with this persistence are unclear. Suppression of viral infections can be mediated by CD8 T cells that detect infected cells via their T cell receptor and the CD8 co-receptor. However, CD8 T cells exhibit signs of functional exhaustion in untreated HIV infection that may not be fully reversed under ART. To investigate whether persistent KS under ART was associated with phenotypic and functional perturbations of CD8 T cells, we performed a cross-sectional study comparing HIV-infected individuals with persistent KS under effective ART (HIV+ KS+) to HIV-infected individuals receiving effective ART with no documented history of KS (HIV+ KS^neg^). A subset of T cells with low cell surface expression of CD8 (“CD8^dim^ T cells”) was expanded in HIV+ KS+ compared with HIV+ KS^neg^ participants. Relative to CD8^bright^ T cells, CD8^dim^ T cells exhibited signs of senescence (CD57) and mitochondrial perturbations (PGC-1α, MitoTracker) ex vivo. Mitochondrial activity (MitoTracker) was also reduced in proliferating CD8^dim^ T cells. These findings indicate that an expanded CD8^dim^ T cell population displaying features of senescence and mitochondrial dysfunction is associated with KS persistence under ART. CD8 co-receptor down-modulation may be symptomatic of ongoing disease.

## Introduction

Kaposi’s sarcoma (KS), a cancer of epithelial and endothelial cells characterized by dark plaques and nodules, is a common AIDS-related morbidity in individuals with untreated HIV infection ^[1]^. However, while the etiologic agent of KS, Kaposi’s sarcoma-associated herpesvirus (KSHV), generates a lifelong infection, it rarely causes disease in immunocompetent individuals. The introduction of HIV antiretroviral therapy (ART), and resulting immune recovery in treated individuals, has been accompanied by a steep decline in HIV-associated KS cases ^[2, 3]^. However, a minority of HIV-infected KSHV-seropositive individuals experience persistent KS despite durable HIV suppression and CD4 T cell recovery under ART ^[4-6]^. The underlying causes of this failure to achieve KS remission are not currently understood. The recent discovery that latent KSHV can be reactivated by proteins from SARS-CoV-2 and some anti-COVID-19 drugs further underscores the need to better understand the control and pathogenesis of KS ^[7]^.

CD8 T cells are major mediators of anti-viral immunity, detecting virus-infected cells via the T cell receptor (TCR) and CD8-co-receptor. KSHV-infected individuals harbor CD8 T cells capable of secreting antiviral cytokines and killing cells expressing KSHV antigens in vitro ^[8-14]^. These KSHV-specific CD8 T cells are detected at higher frequencies in KSHV-seropositive individuals who do not have KS compared with individuals with active disease ^[14]^. Collectively, these observations suggest that in immunocompetent individuals, CD8 T cells may play a lifelong role in preventing KSHV from causing disease. However, during untreated progressive HIV infection, T cells exhibit signs of functional exhaustion that may not be fully reversed by ART ^[15-19]^. Notably, these defects include metabolic perturbations such as impaired mitochondrial oxidative phosphorylation and increased reliance on glycolysis ^[20, 21]^. Mitochondrial metabolism is crucial for lasting CD8 T cell control of viral infections: while effector T cells upregulate glycolysis to rapidly generate ATP, memory T cells use mitochondrial oxidative phosphorylation to support their long-term persistence ^[22-26]^. Indeed, loss of mitochondrial mass is associated with senescence, a state where T cells lose proliferative capacity ^[27]^. Defects in CD8 T cell metabolic fitness in HIV-infected individuals on ART could be particularly detrimental in the tumor microenvironment, where rapidly proliferating malignant cells create an environment of hypoxia and mitochondrial stress ^[28-30]^.

We investigated the possibility that altered CD8 T cell phenotype and metabolism could be associated with persistent KS in HIV-infected individuals on ART by comparing HIV-infected individuals with and without KS. We observed an elevated frequency of CD8^dim^ T cells in individuals with HIV-associated KS. These cells expressed elevated levels of the senescence marker CD57, lower levels of the mitochondrial master-regulator PGC-1α, and exhibited reduced mitochondrial activity. Persistent KS is therefore associated with the expansion of a subset of CD8 T cells with metabolic hallmarks of senescence.

## Materials and Methods

### Study participants

HIV-1-infected participants with biopsy-confirmed KS (“HIV+ KS+”) were recruited from primary care practices in San Francisco and the adjacent counties, UCLA-related primary care clinics, and the Study of the Consequences of the Protease Inhibitor Era (SCOPE). Participants had received ART (including protease inhibitors, NNRTIs, and early integrase inhibitors) and maintained plasma viral loads <75 copies/ml for ≥2 years. CD4 T cell counts were ≥340/µl. All participants had stage 1 tumors according to ACTG criteria (tumor confined to skin and/or lymph nodes and/or minimal oral disease) ^[31]^. The study was approved by the Institutional Review Board of the University of California, San Francisco (approval no. 10-02850). HIV-1-infected participants with no documented history of KS (“HIV+ KS^neg^”) were recruited from the UNC HIV Clinical Trials Unit. Participants had received ART for ≥2 years and maintained plasma viral loads <50 copies/ml and CD4 T cell counts >300/µl for ≥6 months. Participant characteristics are detailed in Table 1. Initial collection of UNC samples was approved by the UNC Institutional Review Board (ethics numbers 11-0228; 14-0741; and 15-1626). Retrospective use of all samples was approved by the UNC Institutional Review Board (ethics number 17-2415).

**Table 1:**
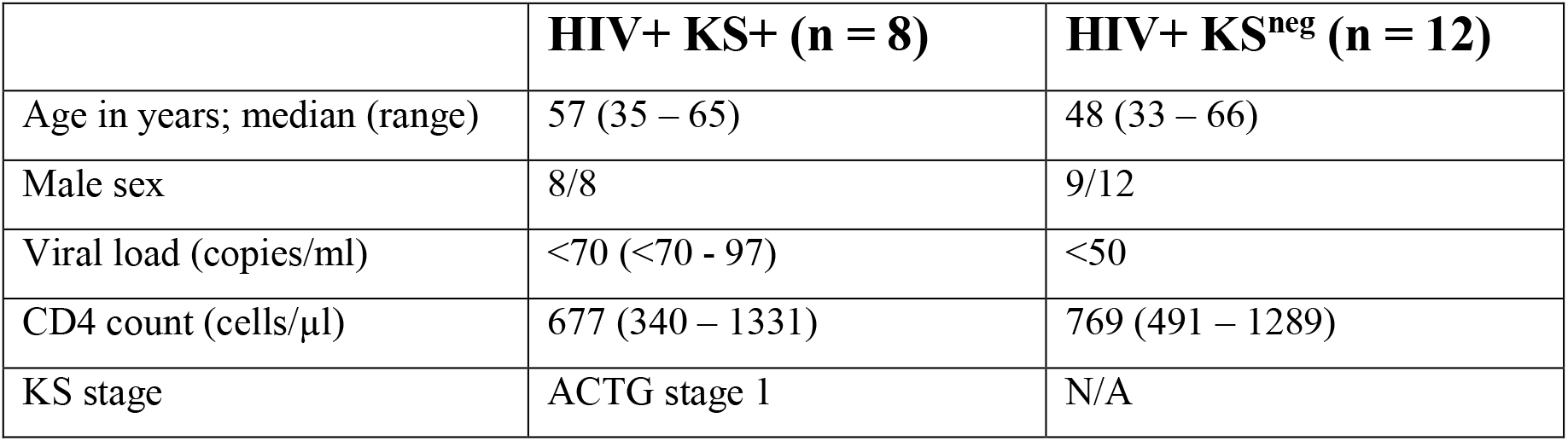
Participant Characteristics.

|  | **HIV+ KS<sup>+</sup> (n = 8)** | **HIV+ KS<sup>neg</sup> (n = 12)** |
| --- | --- | --- |
| Age in years; median (range) | 57 (35 – 65) | 48 (33 – 66) |
| Male sex | 8/8 | 9/12 |
| Viral load (copies/ml) | <70 (<70 - 97) | <50 |
| CD4 count (cells/μl) | 677 (340 – 1331) | 769 (491 – 1289) |
| KS stage | ACTG stage 1 | N/A |

### CD8 T cell phenotyping

Cryopreserved peripheral blood mononuclear cells (PBMC) were rested in R10 medium (RPMI 1640 supplemented with 10% fetal bovine serum; penicillin/streptomycin; 2 mM L-glutamine; 10 mM sodium pyruvate; and 10 mM HEPES) at 37°C overnight. PBMC were stained with Zombie NIR viability dye, then cell surface antibodies CD3-PerCP-Cy5.5 (clone UCHT1); CD4-PE-Cy5 (OKT4); CD8-Brilliant Violet 510 (SK1); CD14 (M5E2), CD16 (3G8), CD19 (HIB19), and CD56 (HCD56)-Brilliant Violet 650; CD45RO-PE (UCHL1); and CD57-PE-Dazzle 594 (HNK-1) (all Biolegend). Cells were stained intracellularly with T-bet-Brilliant Violet 421 (4B10; Biolegend); Eomes-eFluor 660 (WD1928; eBioscience); polyclonal anti-PGC-1α antibody (Santa Cruz Biotechnology); and PE-Cy7 secondary antibody. Samples were acquired on an LSRII flow cytometer and analyzed using FlowJo 10 (BD Biosciences). Live lymphocytes were defined by dim staining with Zombie viability dye, forward scatter height vs area (to identify single events), and forward scatter versus side scatter. CD8 T lymphocytes were defined as CD3+ CD4-CD14/16/19/56- and CD8 bright or dim. For phenotypic markers, positive events were gated using fluorescence minus one controls (Supplementary Figure 1).

### CD8 T cell proliferation

Cryopreserved PBMC were rested overnight, then pulsed under rotation with 5 µM carboxyfluorescein succinimidyl ester (CFSE, Biolegend). Staining was quenched with ice-cold R10. Cells were stimulated with vehicle (0.5% DMSO) or 3 µg/ml phytohaemagglutinin (PHA, Sigma) for 5 days at 37°C. PBMC were stained with Zombie NIR; CD3-PE-Cy7; CD4-BV421; CD8-BV510; CD14, CD16, CD19, and CD56-BV650 (clones as previously; Biolegend). To assess mitochondrial polarization, cells were stained with 25 nM MitoTracker® Deep Red (MTDR, Molecular Probes) at 37°C. Cells were acquired on an LSR Fortessa and analyzed using FlowJo 10 and Modfit LT 4 (Verity Software House). Proliferation was assessed using proliferation index, defined as the mean number of proliferative cycles undergone by each proliferating cell ^[32]^. MTDR^high^ cells were gated using a previously described method ^[33]^. Briefly, after excluding outliers (the brightest and dimmest 0.1% of events), the fluorescence intensities of the brightest and dimmest cells were used to calculate the fluorescence range (brightest – dimmest). Cells that fell within the top 90% of this range were considered MTDR^high^ (Supplementary Figure 2).

### Statistical analysis

Data were analyzed using GraphPad Prism version 8. Between-group differences were analyzed using an exact, two-sided Mann-Whitney U test. Within-individual differences between CD8^bright^ and CD8^dim^ T cells were analyzed using an exact, two-sided Wilcoxon signed-rank test. The monotonic (strictly increasing or decreasing) relationship between variables such as CD8^dim^ percentage and proliferation index was assessed using Spearman’s rank correlation coefficient.

## Results

### CD8^dim^ T cells are expanded in HIV+ individuals with persistent KS, and exhibit features of senescence

We compared the phenotype of CD8 T cells between HIV-infected individuals with persistent KS under ART (HIV+ KS+) and HIV-infected individuals receiving ART with no documented history of KS (HIV+ KS^neg^). CD8 T cells were defined as CD3+ CD8+ CD4-CD14/CD16/CD56- to exclude NKT cells (Supplementary Figure 1). We observed two populations of CD8 T cells: a CD8^bright^ subset with high cell surface expression of CD8 and a CD8^dim^ subset with lower surface expression (Fig. 1A). CD3ε, which forms part of the TCR complex, was also expressed at a lower level on the surface of CD8^dim^ cells compared with CD8^bright^ cells (median of differences = -4358 MFI; Wilcoxon signed-rank test p = 0.007; Supplementary Figure 3). The CD8^dim^ subset was significantly expanded, as a percentage of total CD8 T cells, in HIV+ KS+ compared with HIV+ KS^neg^ participants (difference of medians = 12.28%; Mann-Whitney test p = 0.0006; Fig. 1B). Since highly differentiated T cells accumulate during chronic untreated infections ^[34, 35]^, we next compared the differentiation state of CD8^bright^ and CD8^dim^ cells within participants. CD57 expression was higher on CD8^dim^ than CD8^bright^ T cells (median of differences = 8%; Wilcoxon signed-rank test p = 0.008), indicating that late-differentiated or senescent cells were overrepresented in the CD8^dim^ population (Fig. 1C and Supplementary Figure 3). Supporting this observation, Eomesodermin (Eomes), a transcription factor expressed in terminal memory cells, was expressed in a higher percentage of CD8^dim^ than CD8^bright^ cells (median of differences = 13.5%; Wilcoxon signed-rank test p = 0.008, Fig. 1D and Supplementary Figure 3) ^[36, 37]^. T-bet, a transcription factor expressed by effector cells, was expressed at similar levels in CD8^bright^ and CD8^dim^ cells (Supplementary Figure 3).

**Figure 1.**
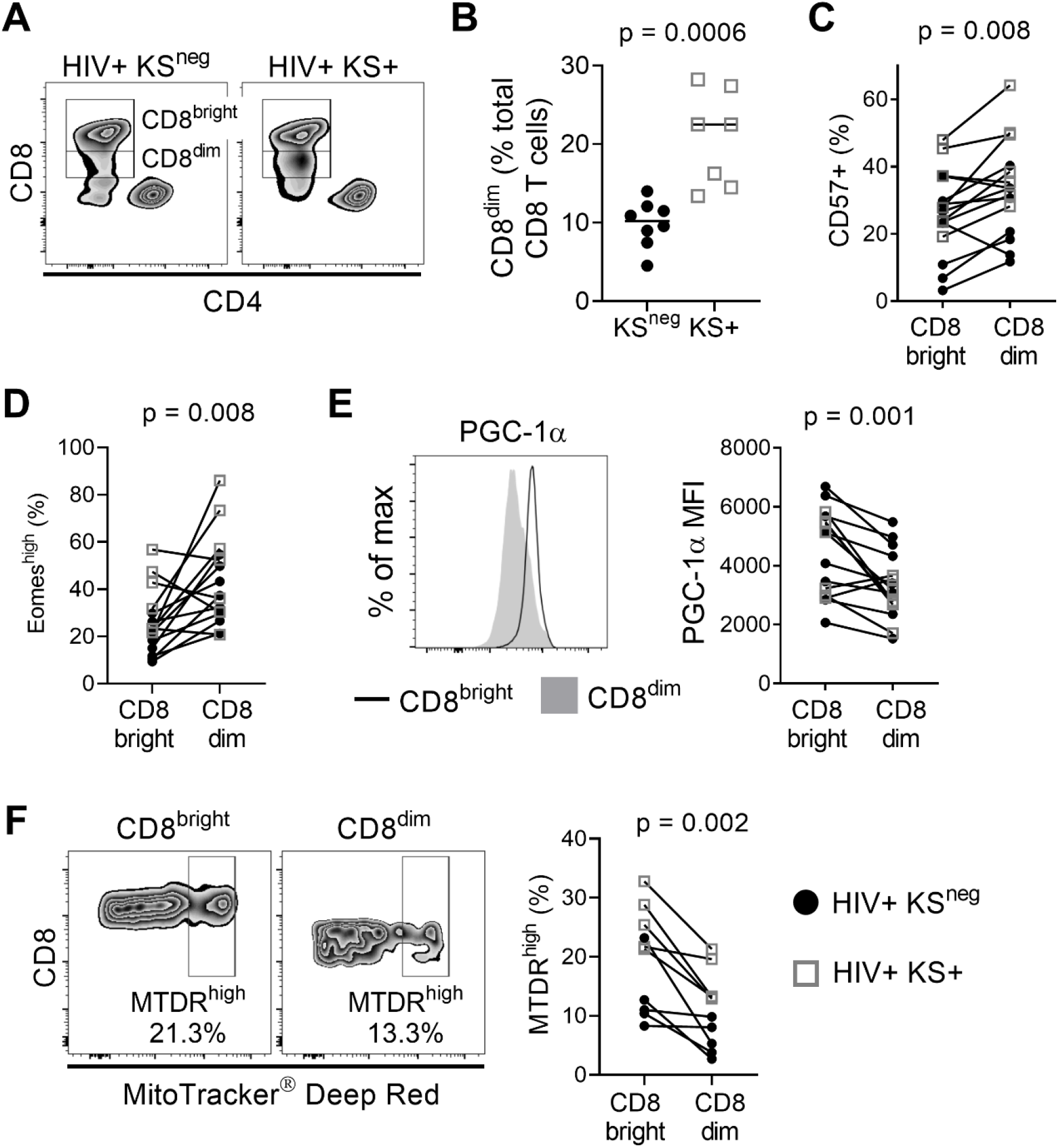
CD8^dim^ cells with low mitochondrial activity are expanded in individuals with persistent KS. A) Representative plots showing CD8^bright^ and CD8^dim^ T cells in a HIV+ KS^neg^ and a HIV+ KS+ participant. B) The frequency of CD8^dim^ T cells (as a percentage of total CD8 T cells) is significantly elevated in HIV+ individuals with persistent KS under ART (KS+; n = 7) compared with the KS^neg^ group (n = 8) (difference of medians= 12.28%; Mann-Whitney test). C) A significantly higher percentage of CD8^dim^ T cells express CD57 compared with CD8^bright^ T cells (median of differences = 8%; Wilcoxon signed-rank test). D) A significantly higher percentage of CD8^dim^ T cells express Eomes compared with CD8^bright^ T cells (median of differences = 13.5%; Wilcoxon signed-rank test). E) Expression of the mitochondrial master regulator PGC-1α is significantly reduced in CD8^dim^ T cells (median of differences = -789 MFI; Wilcoxon signed-rank test). F) The frequency of MitoTracker Deep Red high cells is significantly lower for CD8^dim^ T cells compared with CD8^bright^ T cells (median of differences = -8.99%; Wilcoxon signed-rank test). Gray open squares, HIV+ KS+ participants; black circles, HIV+ KS^neg^ participants.

### CD8^dim^ T cells have an altered mitochondrial phenotype

The development, persistence, and recall function of memory CD8 T cells is highly dependent on mitochondrial metabolism ^[24, 38]^. Compared with CD8^bright^ T cells, expression of the mitochondrial master regulator PGC-1α was significantly reduced in CD8^dim^ T cells (median of differences = -789 MFI; Wilcoxon signed-rank test p = 0.001; Fig. 1E). To further investigate mitochondrial phenotype in CD8^bright^ vs CD8^dim^ T cells, we used MitoTracker® Deep Red (MTDR), which selectively binds actively respiring mitochondria ^[39]^. MTDR^high^ cells were defined using a previously described objective gating strategy (^[33]^; Supplementary Figure 2). The frequency of MTDR^high^ cells was lower in the CD8^dim^ T cell population than CD8^bright^ T cells (median of differences = -8.99%; Wilcoxon signed-rank test p = 0.002; Fig. 1F). These observations indicate that individuals with HIV-associated KS have an expanded population of CD8^dim^ T cells with a highly differentiated/senescent phenotype and reduced mitochondrial activity.

### Mitochondrial activity is reduced in CD8^dim^ proliferating cells

We next examined whether cell surface expression of the CD8 co-receptor was related to replicative capacity and mitochondrial activity in proliferating cells. PBMC were stimulated with the polyclonal stimulus PHA for five days and proliferation was measured by CFSE dilution (Fig. 2A). Proliferative capacity was reported as proliferation index (the mean number of proliferative cycles undergone by each responding cell).

**Figure 2.**
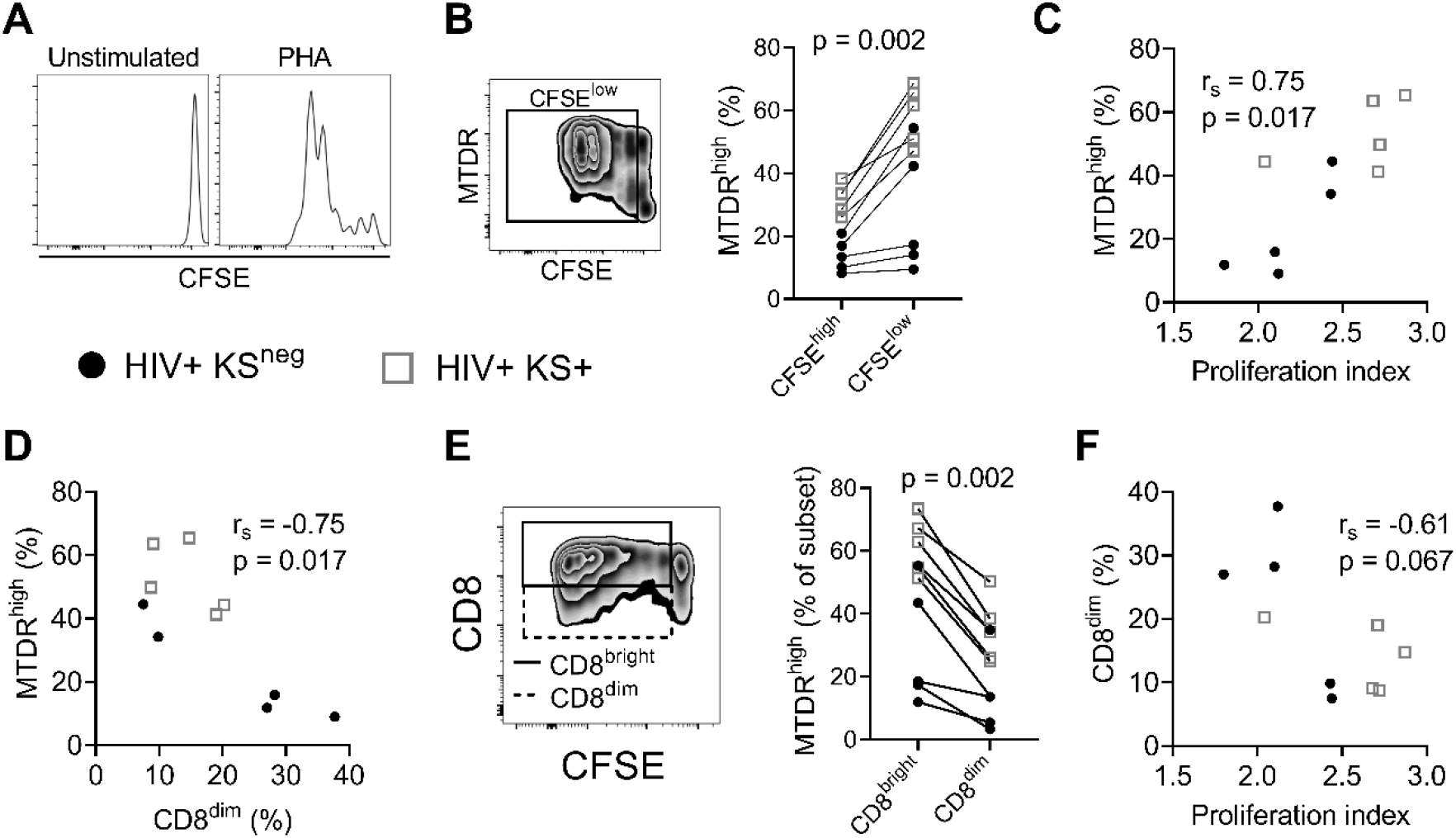
Mitochondrial activity is reduced in CD8^dim^ proliferating cells. A) Histograms showing CFSE dilution in unstimulated and PHA-stimulated CD8 T cells. B) MitoTracker Deep Red (MTDR) fluorescence in PHA-stimulated proliferating (CFSE^low^) cells. A significantly higher percentage of proliferating (CFSE^low^) CD8 T cells are MTDR^high^ compared with non-proliferating (CFSE^high^) CD8 T cells, indicating that proliferating cells have higher mitochondrial activity (n = 10; median of differences = 23.25%; Wilcoxon signed-rank test). C) Positive association between mitochondrial activity (% MTDR^high^) of all CD8 T cells in the culture and the proliferation index of proliferating cells in response to PHA stimulation (Spearman correlation). D) Negative association between mitochondrial activity and the frequency of CD8^dim^ T cells following stimulation with PHA (Spearman correlation). E) The frequency of MTDR^high^ cells, comparing CD8^bright^ and CD8^dim^ proliferating (CFSE^low^) cells. CD8^dim^ proliferating cells are significantly less likely to be MTDR^high^, indicating lower mitochondrial activity (median of differences = -23.35%; Wilcoxon signed-rank test). F) Proliferation index of PHA-stimulated CD8 T cells versus the frequency of CD8^dim^ T cells in the culture (Spearman correlation). Gray open squares, HIV+ KS+ participants; black circles, HIV+ KS^neg^ participants.

CD8 T cells that had proliferated (CFSE^low^) exhibited greater mitochondrial activity (% of cells MTDR^high^) than non-proliferating (CFSE^high^) cells (median of differences = 23.25%; Wilcoxon signed-rank test p = 0.002; Fig. 2B). There was a strong positive correlation between mitochondrial activity of all CD8 T cells and proliferation index at day five, demonstrating the importance of mitochondrial respiration to CD8 T cell proliferation (rs = 0.75, p = 0.017; Fig. 2C) ^[26]^. Conversely, there was a strong negative correlation between mitochondrial activity of all CD8 T cells and the percentage of CD8^dim^ T cells in the culture at five days (rs = -0.75, p = 0.017; Fig. 2D), suggesting that low CD8 expression is associated with reduced mitochondrial activity. Further supporting this hypothesis, CD8^dim^ cells that had proliferated exhibited lower mitochondrial activity (% MTDR^high^) than CD8^bright^ cells that had proliferated (median of differences = -23.35%; Wilcoxon signed-rank test p = 0.002; Fig. 2E). There was also evidence of a moderate negative correlation between the percentage of CD8^dim^ T cells in the culture and proliferation index (rs = -0.61, p = 0.067; Fig. 2F). Collectively these results indicate that low surface expression of CD8 is associated with reduced mitochondrial respiration and replicative capacity of proliferating CD8 T cells.

## Discussion

The underlying causes of persistent KS in a minority of HIV-infected individuals with durable viral suppression and CD4 T cell recovery under ART are unknown. Here, we have identified an expanded population of CD8^dim^ T cells with phenotypic characteristics of senescence and mitochondrial dysfunction in these individuals.

The CD8 co-receptor amplifies signals through the TCR ^[40, 41]^. Following antigenic stimulation (e.g. by a virus-infected cell), CD8, along with the TCR, is downregulated from the cell surface, possibly to limit the strength or duration of signaling ^[42-44]^. This suggests that the expansion of CD8^dim^ T cells may be a response to high and/or persistent antigen stimulation. Supporting this hypothesis, elevated frequencies of CD8^dim^ T cells have been reported during acute HIV infection, and in children exposed to a high cumulative pathogen burden during the first years of life ^[45, 46]^. Our observation that CD8^dim^ T cells are also expanded in individuals with persistent KS under ART supports the notion that CD8 downregulation is a general phenomenon in settings of unresolved infection.

Chronic viral infections are also associated with CD8 T cell terminal differentiation and/or senescence^[47]^. Senescent CD8 T cells exhibit reduced expression of PGC-1α, the master-regulator of mitochondrial biogenesis, and lower mitochondrial activity ^[27, 48]^. Conversely, forced expression of PGC-1α promotes robust CD8 T cell memory responses ^[49]^. These observations underscore the importance of mitochondrial respiration for long-term CD8 T cell anti-viral function. In the setting of persistent KS, we observed that the expanded CD8^dim^ T cell population expressed high levels of Eomesodermin and CD57, proteins respectively associated with terminal differentiation and senescence, and had reduced expression of PGC-1α. CD8^dim^ cells also contained fewer respiring mitochondria both *ex vivo* and when proliferating. Our data suggest that mitochondrial dysfunction may underlie the accumulation of senescent CD8 T cells that has previously been reported in KS ^[34]^; however, this hypothesis must be tested in future studies. It is unclear from our current data whether CD8 expression directly regulates mitochondrial activity in CD8 T cells.

A key question is whether the CD8^dim^ T cells we observed are specific for KSHV, for HIV, or for other persistent viral infections such as CMV, which is highly seroprevalent in HIV-infected individuals ^[50]^. It is possible that CD8 down-modulation could be driven by multiple concurrent infections, by ongoing immune activation, or by a combination of factors. In this initial study, we were unable to determine whether the CD8^dim^ T cells we observed were KSHV-specific, as KSHV is a large virus, and immunoprevalent epitopes eliciting responses in a high percentage of seropositive individuals have not yet been identified. The question of the antigen specificity of CD8^dim^ T cells will be the subject of subsequent investigations.

Our work has some limitations. As this was an observational study, we were unable to determine whether the expansion of CD8^dim^ T cells plays a causative role in the failure to control KSHV under ART or is a consequence of this lack of suppression. Due to lack of available tissue, we were unable to assess whether CD8 T cells infiltrating the tumor microenvironment also exhibit a CD8^dim^ phenotype. This question, together with the antigen specificity of CD8^dim^ T cells and a direct examination of their functional profile, is the subject of ongoing investigations. If KSHV-specific CD8 T cells infiltrating the tumor microenvironment express low levels of CD8 and exhibit reduced mitochondrial activity, this will have important implications for immunotherapeutic approaches to KS treatment.

## Data Availability

All data produced in the present study are available upon reasonable request to the authors.

## Abbreviations used in this article

ART: (HIV) Anti-retroviral therapy
CFSE: carboxyfluorescein succinimidyl ester
KS: Kaposi’s sarcoma
KSHV: Kaposi’s sarcoma-associated herpesvirus, also known as HHV-8
MTDR: MitoTracker® Deep Red
PBMC: Peripheral blood mononuclear cells
PHA: Phytohaemagglutinin
TCR: T cell receptor

## Author contributions

The manuscript was written by G.T.C., A.M.W., N.P.G., and T.M. G.T.C. and N.P.G. contributed to study design. G.T.C. performed experimentation and collection of data. G.T.C. and A.M.W. performed the statistical analyses. G.T.C. acquired funding for the study. T.M. facilitated participant recruitment and sample collection. All authors provided review of the final manuscript. We thank Joann Kuruc and Cynthia Gay for their work recruiting participants to UNC cohorts that were retrospectively utilized for this study.

## Conflicts of interest

There are no conflicts of interest.

## Notes

Funding: This study was supported by a Young Investigator Pilot Award awarded to G.T.C. by the AIDS and Cancer Specimen Resource, funded by the National Cancer Institute (UM1 CA181255). Initial sample collection was supported by NIH grants U01 AI117844, U01 AI095052, and R01 HL132791. The UNC Flow Cytometry Core Facility is supported in part by P30 CA016086 Cancer Center Core Support Grant to the UNC Lineberger Comprehensive Cancer Center and by the Center for AIDS Research award number 5P30AI050410. Statistical expertise was provided by the University of North Carolina at Chapel Hill Center for AIDS Research, an NIH funded program P30 AI050410. The content is solely the responsibility of the authors and does not necessarily represent the official views of the National Institutes of Health.

### Competing Interest Statement

The authors have declared no competing interest.

### Funding Statement

This study was supported by a Young Investigator Pilot Award awarded to G.T.C. by the AIDS and Cancer Specimen Resource, funded by the National Cancer Institute (UM1 CA181255). Initial sample collection was supported by NIH grants U01 AI117844, U01 AI095052, and R01 HL132791. The UNC Flow Cytometry Core Facility is supported in part by P30 CA016086 Cancer Center Core Support Grant to the UNC Lineberger Comprehensive Cancer Center and by the Center for AIDS Research award number 5P30AI050410. Statistical expertise was provided by the University of North Carolina at Chapel Hill Center for AIDS Research, an NIH funded program P30 AI050410. The content is solely the responsibility of the authors and does not necessarily represent the official views of the National Institutes of Health.

### Author Declarations

The study of participants with KS was approved by the Institutional Review Board of the University of California, San Francisco (approval no. 10-02850). Initial collection of UNC samples was approved by the UNC Institutional Review Board (ethics numbers 11-0228; 14-0741; and 15-1626). Retrospective use of all samples was approved by the UNC Institutional Review Board (ethics number 17-2415).

