## Supplementary materials for "An expanded population of CD8^dim^ T cells with features of mitochondrial dysfunction and senescence is associated with persistent HIV-associated Kaposi’s sarcoma under ART"

### Supplementary Figures

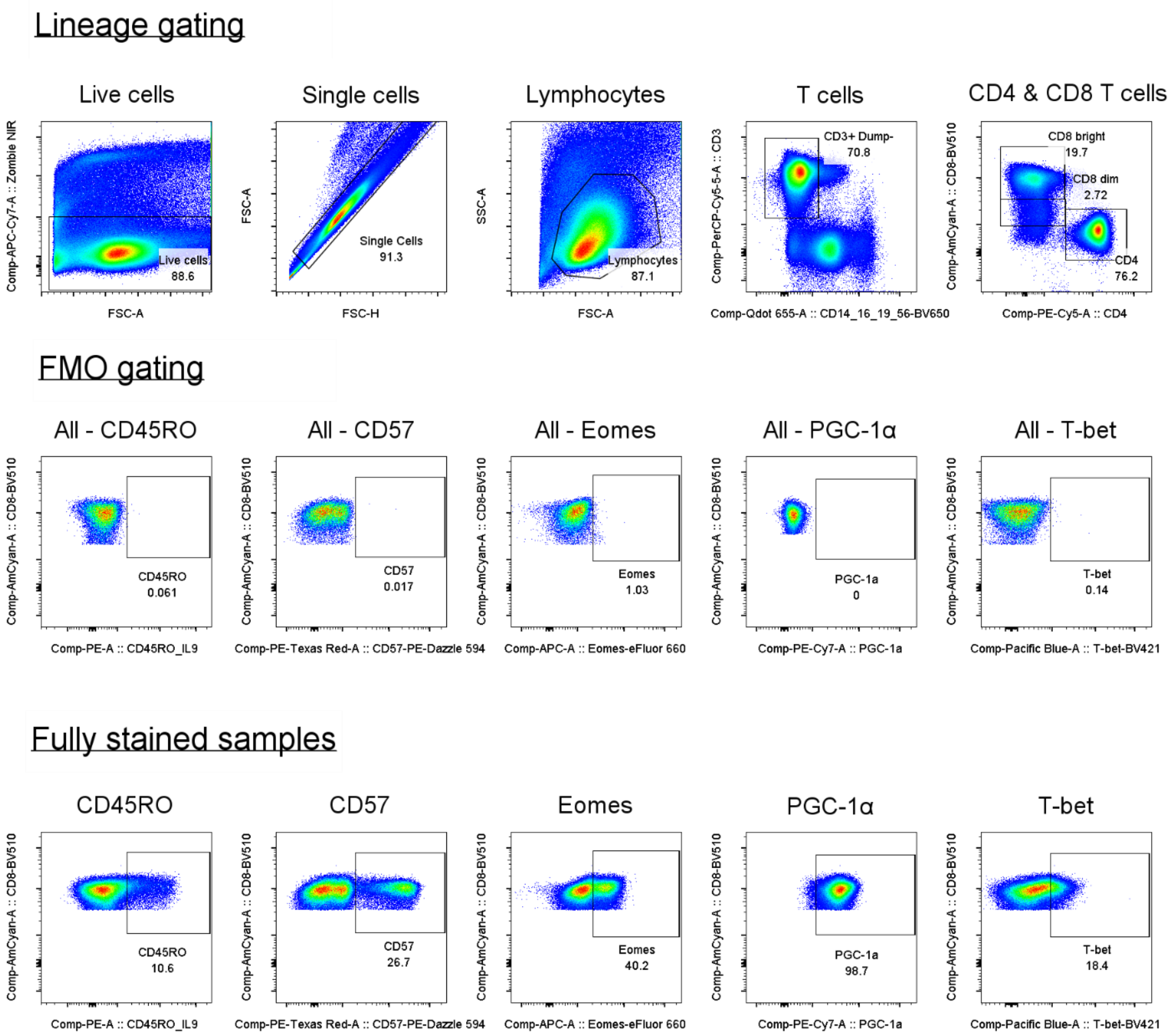


**Supplementary Figure 1.** Gating strategy used to identify CD8 bright and dim T cells (top row) and their expression of phenotypic markers (bottom row). Gates were set using fluorescence minus one (FMO) controls (middle row).

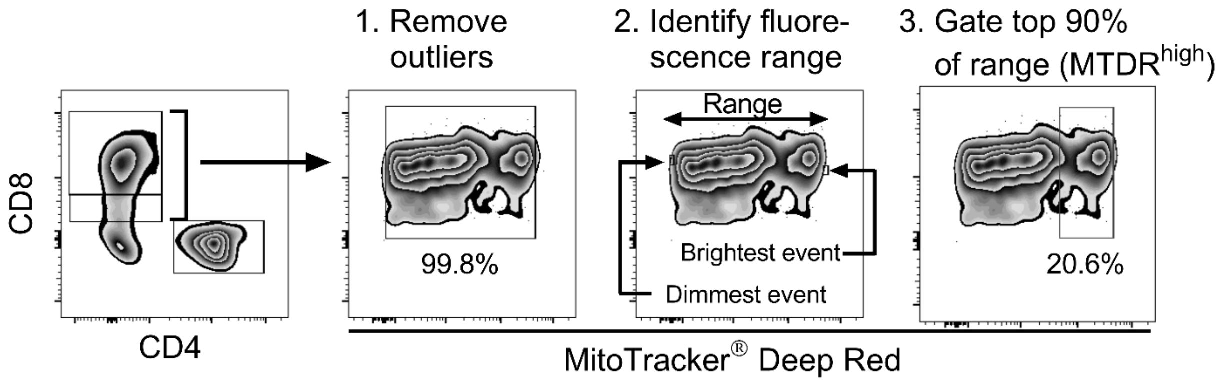


**Supplementary Figure 2.** Gating strategy used to delineate MitoTracker Deep Red (MTDR) "high" CD8 T cells. After excluding outliers (top and bottom 0.1%), the fluorescence intensity of the brightest and dimmest cells indicated the fluorescence range. Cells that fell within the top 90% of this fluorescence range were considered $MTDR^{high}$. (Note: plots use bi-exponential scale.)

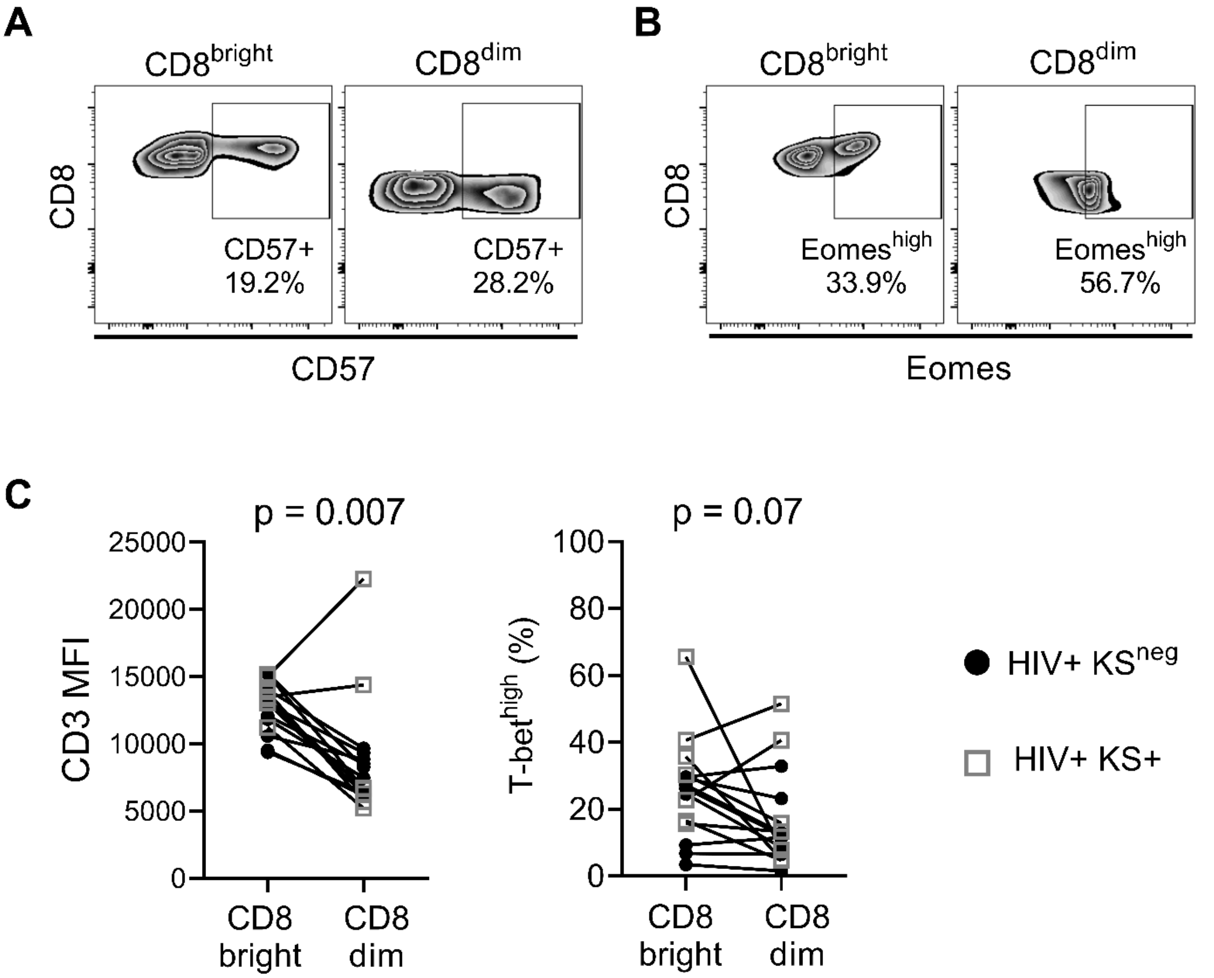


**Supplementary Figure 3.** A) Representative plots of CD57 expression on CD8$^{bright}$ and CD8$^{dim}$ T cells. B) Representative plots of Eomes expression on CD8$^{bright}$ and CD8$^{dim}$ T cells. C) CD3 and T-bet expression, comparing CD8$^{bright}$ and CD8$^{dim}$ T cells from HIV+ KS+ (gray open squares) and HIV+ KS$^{neg}$ (black circles) participants.
